# Disability associated with failure to seek medical care among Venezuelan immigrants in Peru

**DOI:** 10.1101/2023.07.11.23292412

**Authors:** Mercedes Miranda-Tueros, Sonny Sthefanie Velarde-Meza, J. Jhonnel Alarco

## Abstract

**Background:** Currently, there are more than six million Venezuelan immigrants worldwide. This study aimed to estimate the association between disability and failure to seek medical care among Venezuelan immigrants in Peru.

**Methods:** A cross-sectional study was conducted using secondary data of the Encuesta Dirigida a la Población Venezolana que Reside en el País (ENPOVE) 2018. We developed four Poisson regression models and calculated prevalence ratios (PR) with their 95% confidence intervals (95% CI).

**Results:** After adjusting for multiple confounding variables, we found that immigrants who reported having only one type of disability were 78% more likely not to seek medical care compared with immigrants without disability (PR = 1.78; 95% CI 1.15–2.76).

**Conclusions:** Venezuelan immigrants with disabilities in Peru seek less medical care than those without disabilities.

## Introduction

Worldwide, approximately 6.7 million persons with disabilities of 45.2 million immigrants are forcibly displaced because of persecution, conflict, generalized violence, and human rights violations [1].

According to updated figures up to September 2022 from the United Nations Refugee Agency’s Coordination Platform for Refugees and Migrants in Venezuela (R4V), the number of Venezuelan immigrants exceeds six million worldwide (6,133,473) [2].

Countries such as Colombia, Ecuador, Peru, and Chile are the primary recipients of these immigrants [3]. In addition, migrant populations experience several challenges in host countries, such as cultural and language barriers, stigma, discrimination, occupational hazards, social exclusion, and poor accessibility to health systems [4]. Thus, people with disabilities experience adverse situations, such as increased health problems and discrimination [5]. Therefore, we can conjecture that immigrants with disabilities would find themselves in very vulnerable situations.

Few population-based studies have evaluated immigrant disability and health care-seeking behaviors, especially in developing countries with substantial health inequities. The few studies were conducted in small groups and using qualitative methodology [6,7]. Therefore, this study aimed to estimate the association between disability and the non-seeking of health care in Venezuelan immigrants in Peru. We hypothesized that Venezuelan immigrants with disabilities would seek less medical care after becoming ill than their peers without disabilities.

## Materials and Methods

### Design and population

A cross-sectional study was conducted using secondary data of the Encuesta Dirigida a la Población Venezolana que Reside en el País (ENPOVE), which was carried out by the Instituto Nacional de Estadística e Informática (INEI) of Peru in 2018. This survey was applied in six cities in Peru, namely, Tumbes, La Libertad, Arequipa, Cusco, Lima, and Callao, which were selected for having a higher proportion of Venezuelan immigrants. The ENPOVE was conducted from November 20 to December 31, 2018 [8].

### Source of data

ENPOVE aimed to provide demographic, social, migration, discrimination, violence, housing, essential services, and household equipment information on the Venezuelan population in Peru. The ENPOVE included every household member aged ≥12 years; when the participant was younger than 12 years old, the head of household provided the data. The sample size calculated by INEI was 3611 residences, including 3697 households and 9847 Venezuelan immigrants. This sample provided representative results for the study cities [8].

The ENPOVE sampling was probabilistic, stratified, and conducted independently in each region. The sampling had two units; the first unit was the block that corresponds to the geographic area of each selected dwelling. The second was the residence with at least one Venezuelan immigrant. The research unit consisted of residences, households, and Venezuelan nationals. Relevant information was obtained by direct interview and was collected on a digital tablet, containing the questionnaire, which allowed direct transmission of data. The interviewers visited the selected households until all member information was complete [8].

ENPOVE defined disability as “a permanent physical, mental, intellectual or sensory impairment that prevents the adequate performance of daily activities or the full exercise of rights” [8].

For this study, we included the records of Venezuelan immigrants of both sexes who reported having an illness after they arrived in Peru and those who were working (aged ≥14 years) [9]. Records with missing data were excluded.

### Variables

The dependent variable was not seeking medical attention after illness, symptoms, or relapse after they arrived in Peru, according to the question, “Where did you go to consult for this illness, symptom, discomfort, and/or accident?.” The response alternatives were as follows: MINSA (Ministry of Health), EsSalud (Social Health Insurance), outpatient clinic, private clinic, pharmacy or drugstore, self-medicated, others, and did not seek care. Not seeking medical care was considered positive when immigrants responded only to the alternative “not seeking care.” Finally, this variable assumed two categories: do not seek medical care or seek medical care. Other studies have measured this variable similarly [10].

The independent variable was disability, according to the following questions: “Do you have permanent limitations, to move or walk, or to use arms or legs? Can you see even when wearing glasses? Can you speak or communicate even when using sign language or others? Can you hear even when using hearing aids? Can you understand or learn (concentrate and remember)? Can you relate to others’ thoughts, feelings, emotions, or behaviors?.” The immigrant would have a disability if he/she answered yes to at least one of the above questions. These questions correspond to the Washington group’s shortlist measurement of disability [11]. Finally, this variable assumed three categories: no disability, only one type of disability, and two or more types of disability. Other studies have measured this variable in a similar way [5].

We included sociodemographic variables such as sex (male and female), age (age groups), education (no education/preschool/primary, secondary, or higher), and marital status (married, cohabiting, widowed/divorced/separated, or single). We also included variables related to not seeking medical care, such as health insurance (no or yes) [12], chronic illness (no or yes) [13], current job (no or yes) [14], institutional help [“Since you arrived in Peru, have you received any institutional help?” (no or yes)] [15], ethnic self-identification (mestizo, Afro-descendant, white, indigenous, or others/do not know) [16], sexual orientation (heterosexual or lesbian, gay, transsexual, bisexual, and intersexual [LGTBI]) [17], and perception of discrimination because of nationality [“Have you felt discriminated against for being a Venezuelan since you arrived in Peru?” (no or yes)] [18], and length of stay in Peru (months).

### Statistical analysis

The databases were downloaded in their original format (*sav* extension) from the INEI web page [19] and imported into Stata MP version 16 for Windows®, where they were merged and analyzed. Frequencies and weighted percentages with their 95% confidence intervals (95% CI) were calculated. Differences in health care-seeking behavior were analyzed with the chi-square test (corrected for survey design) for categorical variables and with the Wald test for numerical variables. To estimate the magnitude of the association, we used generalized linear models of the Poisson family with logarithmic linkage and calculated prevalence ratios (PR) with their 95% CIs. Four regression models were developed according to epidemiological criteria, including the primary association. The first model showed a crude association. The second model was adjusted for sociodemographic variables, the third model was adjusted for variables related to not seeking medical care, and the fourth model was adjusted for all variables included in the study. A value of *p* < 0.05 was accepted as significant. Possible multicollinearity was assessed with the variance inflation factor (VIF).

## Results

The initial sample had 8836 records. Of these, 4592 immigrants who did not present any diseases after they arrived in Peru and 431 who were under 14 years of age were excluded, leaving 3813 records for the final analysis (Figure 1).

**Figure 1.**
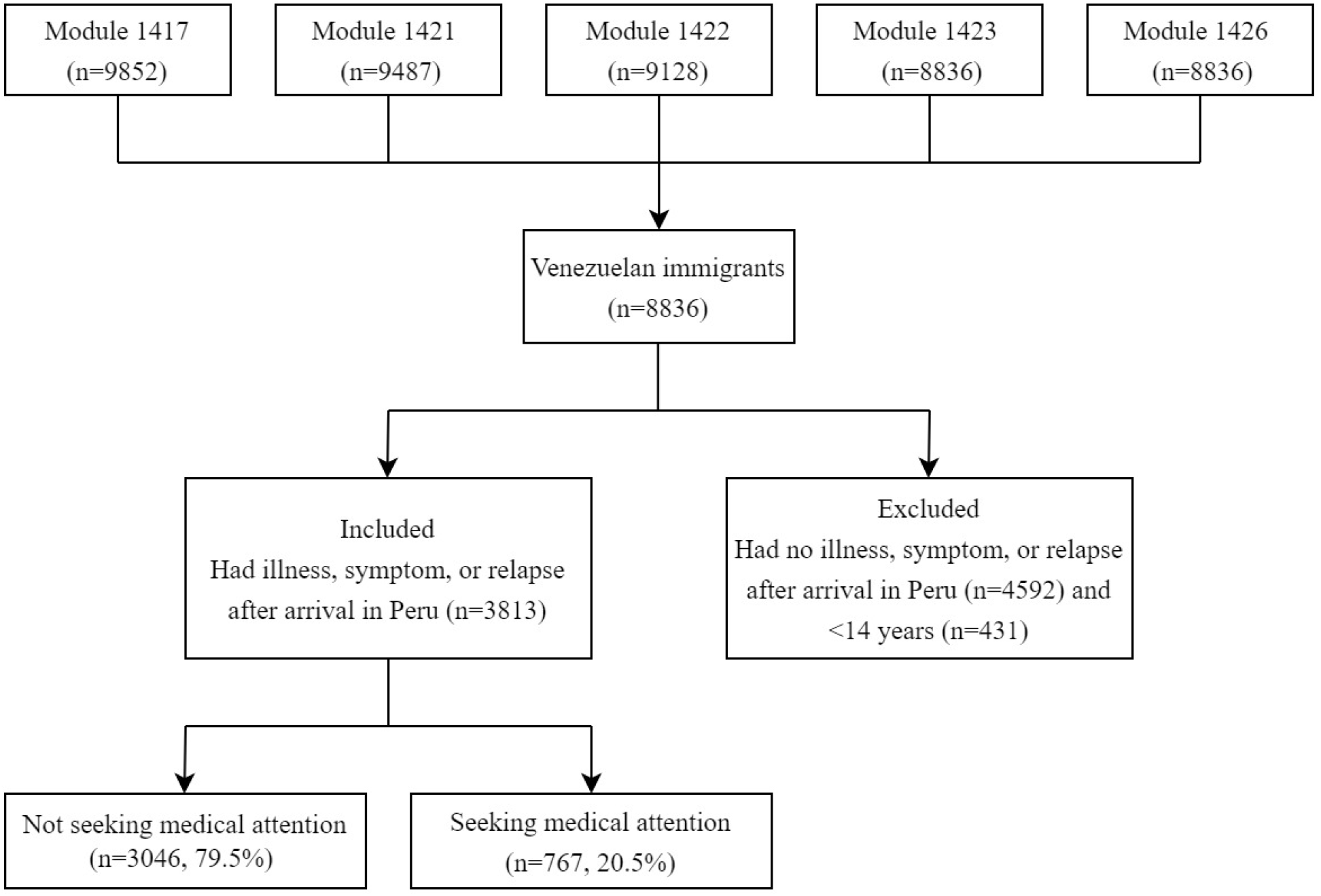
Selection flow chart of Venezuelan immigrants in Peru included in the study.

In total, 2.1% (n = 80) reported having a disability, of which 85% (n = 68) had only one type of disability and 15% (n = 12) had two or more types of disability. In addition, 20.5% (n = 767) did not seek medical attention when they experienced illness, symptom, or relapse after they arrived in Peru. Most of the Venezuelan immigrants were young (18–29 years old), male, single, and with higher education. Likewise, most of them had no chronic diseases, had no health insurance, had a job at the time of the interview, had not received any institutional help after their arrival in Peru, did not perceive discrimination because of their nationality, were heterosexual, and self-defined as mestizo (Table 1).

**Table 1.**
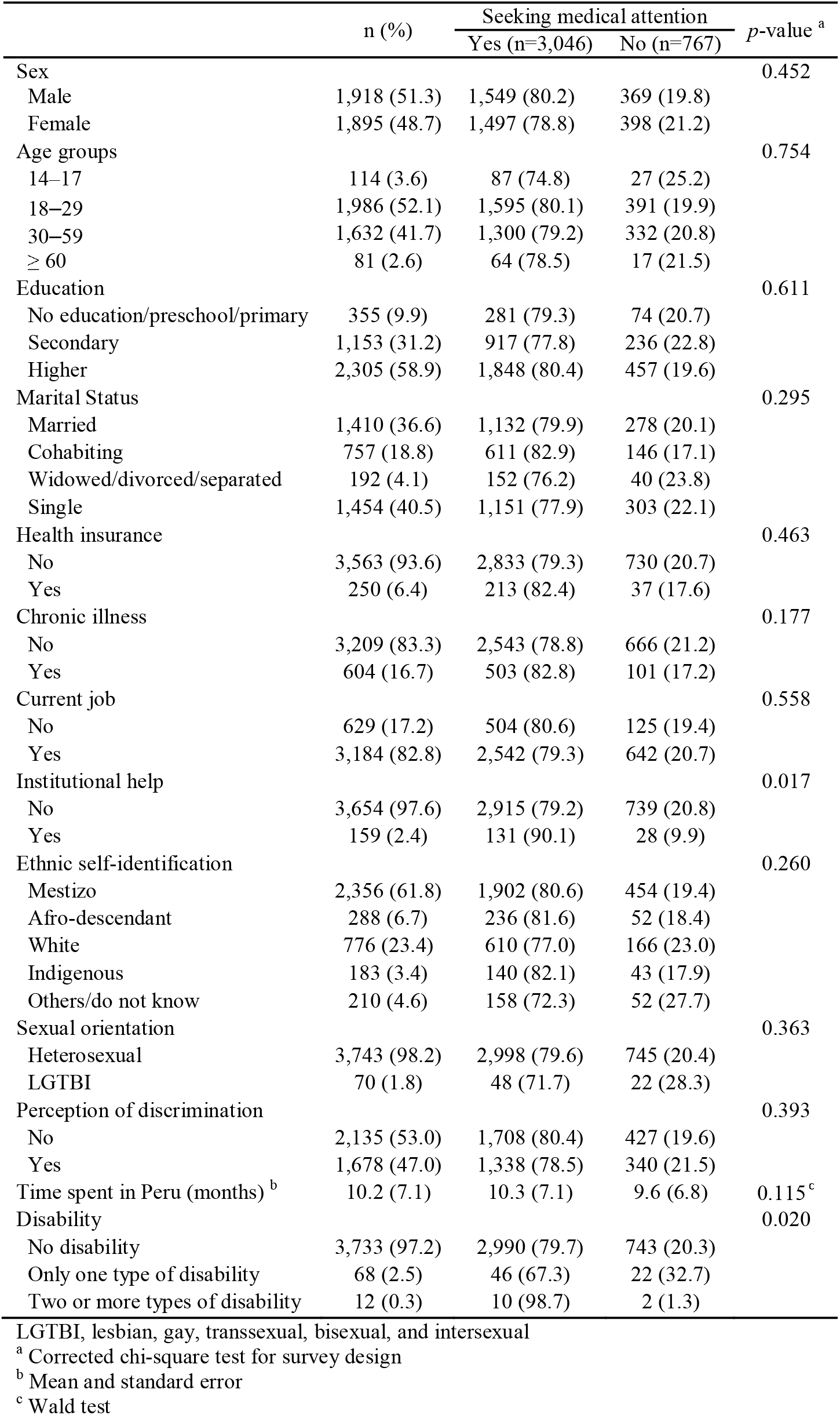
Characteristics and differences in medical attention-seeking among Venezuelan immigrants in Peru.

|  | n (%) | Seeking medical attention |  | *p*-value <sup>a</sup> |
| --- | --- | --- | --- | --- |
|  |  | Yes (n=3,046) | No (n=767) |  |
| Sex |  |  |  | 0.452 |
| Male | 1,918 (51.3) | 1,549 (80.2) | 369 (19.8) |  |
| Female | 1,895 (48.7) | 1,497 (78.8) | 398 (21.2) |  |
| Age groups |  |  |  | 0.754 |
| 14–17 | 114 (3.6) | 87 (74.8) | 27 (25.2) |  |
| 18–29 | 1,986 (52.1) | 1,595 (80.1) | 391 (19.9) |  |
| 30–59 | 1,632 (41.7) | 1,300 (79.2) | 332 (20.8) |  |
| ≥ 60 | 81 (2.6) | 64 (78.5) | 17 (21.5) |  |
| Education |  |  |  | 0.611 |
| No education/preschool/primary | 355 (9.9) | 281 (79.3) | 74 (20.7) |  |
| Secondary | 1,153 (31.2) | 917 (77.8) | 236 (22.8) |  |
| Higher | 2,305 (58.9) | 1,848 (80.4) | 457 (19.6) |  |
| Marital Status |  |  |  | 0.295 |
| Married | 1,410 (36.6) | 1,132 (79.9) | 278 (20.1) |  |
| Cohabiting | 757 (18.8) | 611 (82.9) | 146 (17.1) |  |
| Widowed/divorced/separated | 192 (4.1) | 152 (76.2) | 40 (23.8) |  |
| Single | 1,454 (40.5) | 1,151 (77.9) | 303 (22.1) |  |
| Health insurance |  |  |  | 0.463 |
| No | 3,563 (93.6) | 2,833 (79.3) | 730 (20.7) |  |
| Yes | 250 (6.4) | 213 (82.4) | 37 (17.6) |  |
| Chronic illness |  |  |  | 0.177 |
| No | 3,209 (83.3) | 2,543 (78.8) | 666 (21.2) |  |
| Yes | 604 (16.7) | 503 (82.8) | 101 (17.2) |  |
| Current job |  |  |  | 0.558 |
| No | 629 (17.2) | 504 (80.6) | 125 (19.4) |  |
| Yes | 3,184 (82.8) | 2,542 (79.3) | 642 (20.7) |  |
| Institutional help |  |  |  | 0.017 |
| No | 3,654 (97.6) | 2,915 (79.2) | 739 (20.8) |  |
| Yes | 159 (2.4) | 131 (90.1) | 28 (9.9) |  |
| Ethnic self-identification |  |  |  | 0.260 |
| Mestizo | 2,356 (61.8) | 1,902 (80.6) | 454 (19.4) |  |
| Afro-descendant | 288 (6.7) | 236 (81.6) | 52 (18.4) |  |
| White | 776 (23.4) | 610 (77.0) | 166 (23.0) |  |
| Indigenous | 183 (3.4) | 140 (82.1) | 43 (17.9) |  |
| Others/do not know | 210 (4.6) | 158 (72.3) | 52 (27.7) |  |
| Sexual orientation |  |  |  | 0.363 |
| Heterosexual | 3,743 (98.2) | 2,998 (79.6) | 745 (20.4) |  |
| LGTBI | 70 (1.8) | 48 (71.7) | 22 (28.3) |  |
| Perception of discrimination |  |  |  | 0.393 |
| No | 2,135 (53.0) | 1,708 (80.4) | 427 (19.6) |  |
| Yes | 1,678 (47.0) | 1,338 (78.5) | 340 (21.5) |  |
| Time spent in Peru (months) <sup>b</sup> | 10.2 (7.1) | 10.3 (7.1) | 9.6 (6.8) | 0.115 <sup>c</sup> |
| Disability |  |  |  | 0.020 |
| No disability | 3,733 (97.2) | 2,990 (79.7) | 743 (20.3) |  |
| Only one type of disability | 68 (2.5) | 46 (67.3) | 22 (32.7) |  |
| Two or more types of disability | 12 (0.3) | 10 (98.7) | 2 (1.3) |  |

LGTBI, lesbian, gay, transsexual, bisexual, and intersexual
<sup>a</sup> Corrected chi-square test for survey design
<sup>b</sup> Mean and standard error
<sup>c</sup> Wald test

In the bivariate analysis, when evaluating the differences according to whether or not medical care was sought, only institutional help and disability status presented significant differences. In addition, 32.7% of the people with a type of disability did not seek medical care compared with 20.3% of people without disability who did not seek medical care; this difference was significant (p = 0.020) (Table 1).

In the multivariate analysis, the association between disability and not seeking medical care remained constant and significant, according to the models considered. In model 1 (crude) and model 2 adjusted for sociodemographic variables, the magnitudes were comparable (PR = 1.61; 95% CI 1.04–2.50; PR = 1.63; 95% CI 1.04–2.54, respectively). In model 3, adjusted for variables related to not seeking medical care, an increase in magnitude was observed (PR = 1.78; 95% CI 1.16–2.74). Finally, in model 4, adjusted for all variables, people with only one type of disability were 78% more likely not to seek medical care than people without disability, adjusted for age group, sex, marital status, education, chronic disease, health insurance, current job, institutional help, perception of discrimination, ethnic self-identification, sexual orientation, and time spent in Peru (PR = 1.78; 95% CI 1.15–2.76) (Table 2). No evidence of multicollinearity was found among the exposure variables in the final model (VIF < 10).

**Table 2.** Association between disability and failure to seek medical care according to the proposed models.

|  | Model 1<br>PR (95% IC) | Model 2<br>PR (95% IC) | Model 3<br>PR (95% IC) | Model 4<br>PR (95% IC) |
| --- | --- | --- | --- | --- |
| Disability |  |  |  |  |
| No disability | Ref | Ref | Ref | Ref |
| Only one type of disability | 1.61 (1.04 – 2.50) | 1.63 (1.04 – 2.54) | 1.78 (1.16 – 2.74) | 1.78 (1.15 – 2.76) |
| Two or more types of disability | 0.06 (0.10 – 0.38) | 0.05 (0.008 – 0.34) | 0.06 (0.01 – 0.42) | 0.05 (0.01 – 0.37) |
| Sex |  |  |  |  |
| Male |  | Ref |  | Ref |
| Female |  | 1.08 (0.90 – 1.31) |  | 1.10 (0.92 – 1.32) |
| Age groups |  |  |  |  |
| 14–17 |  | 1.18 (0.74 – 1.89) |  | 1.24 (0.75 – 2.06) |
| 18–29 |  | Ref |  | Ref |
| 30–59 |  | 1.09 (0.88 – 1.35) |  | 1.12 (0.90 – 1.40) |
| ≥ 60 |  | 1.14 (0.58 – 2.23) |  | 1.34 (0.65 – 2.74) |
| Education |  |  |  |  |
| No education/preschool/primary |  | 1.09 (0.86 – 1.38) |  | 1.06 (0.84 – 1.33) |
| Secondary |  | 1.01 (0.68 – 1.48) |  | 0.98 (0.66 – 1.45) |
| Higher |  | Ref |  | Ref |
| Marital Status |  |  |  |  |
| Married |  | Ref |  | Ref |
| Cohabiting |  | 0.81 (0.60 – 1.10) |  | 0.84 (0.62 – 1.15) |
| Widowed/divorced/separated |  | 1.20 (0.74 – 1.95) |  | 1.24 (0.75 – 2.03) |
| Single |  | 1.09 (0.86 – 1.37) |  | 1.08 (0.85 – 1.37) |
| Health insurance |  |  |  |  |
| No |  |  | Ref | Ref |
| Yes |  |  | 0.95 (0.59 – 1.51) | 0.99 (0.62 – 1.60) |
| Chronic illness |  |  |  |  |
| No |  |  | Ref | Ref |
| Yes |  |  | 0.80 (0.58 – 1.10) | 0.78 (0.56 – 1.09) |
| Current job |  |  |  |  |
| No |  |  | Ref | Ref |
| Yes |  |  | 1.00 (0.80 – 1.27) | 1.02 (0.79 – 1.32) |
| Institutional help |  |  |  |  |
| No |  |  | Ref | Ref |
| Yes |  |  | 0.47 (0.24 – 0.93) | 0.49 (0.24 – 0.97) |
| Ethnic self-identification |  |  |  |  |
| Mestizo |  |  | Ref | Ref |
| Afro-descendant |  |  | 0.92 (0.61 – 1.37) | 0.92 (0.62 – 1.36) |
| White |  |  | 1.18 (0.92 – 1.51) | 1.20 (0.93 – 1.54) |
| Indigenous |  |  | 0.90 (0.55 – 1.47) | 0.91 (0.56 – 1.49) |
| Others/do not know |  |  | 1.40 (0.90 – 2.18) | 1.37 (0.90 – 2.08) |
| Sexual orientation |  |  |  |  |
| Heterosexual |  |  | Ref | Ref |
| LGTBI |  |  | 1.49 (0.78 – 2.87) | 1.47 (0.75 – 2.88) |
| Perception of discrimination |  |  |  |  |
| No |  |  | Ref | Ref |
| Yes |  |  | 1.10 (0.89 – 1.38) | 1.13 (0.90 – 1.40) |
| Time spent in Peru (months) |  |  | 0.99 (0.97 – 1.00) | 0.99 (0.97 – 1.01) |

PR, prevalence ratio; 95% CI, 95% confidence interval; Ref, reference variable; LGTBI, lesbian, gay, transgender, bisexual, and intersexual
Model 1: raw
Model 2: adjusted for sociodemographic variables (sex, age group, education, and marital status)
Model 3: adjusted for variables related to not seeking medical care (health insurance, chronic illness, current job, institutional help, ethnic self-identification, sexual orientation, perception of discrimination, and time spent in Peru)
Model 4: adjusted for all variables

## Discussion

In this study, we found an association between disability and failure to seek medical care among Venezuelan immigrants in Peru. This association remained constant and significant after adjusting for sociodemographic variables and variables related to not seeking medical care.

The prevalence of disability was 2.1%. Although there is limited information on the situation and prevalence of immigrants with disabilities worldwide [20], some studies have reported higher figures. Based on secondary data from the Survey of Disabilities, Impairments and Health Conditions in Spain, a prevalence of 5% (225,000 people) of immigrants with disabilities, mostly of working age, was estimated [21]. In Chile, a prevalence of 3.55% was reported, almost half the prevalence reported in the Chilean population (6.93%) [4]. The prevalence of disability in Peru, measured with the same ENPOVE questions, was 5.2% [22], almost double that found in Venezuelan immigrants. This difference could be due to the “healthy migrant effect” [4], which refers to newcomers enjoying better health, with a lower probability of reporting chronic or disabling diseases.

The prevalence of not seeking medical care was 20.5%, a lower figure than reported in a similar study conducted in the general population of Peru, which found a prevalence of non-use of health services of 53.9% [23], although with slight modifications in the measurements. However, another study conducted on the LGTBI population in Peru reported a prevalence of 16% of not seeking medical care [24], a figure more in line with our results. In Latin America, prevalence figures vary according to the country, with Colombia and Brazil reporting 33.2% and 25.7%, respectively, in studies conducted on the general population [25]. However, evidence suggests that immigrants use fewer health services than native people. Two systematic reviews, one conducted between 1980 and 2003, which included 37 studies [26], and another conducted between 2013 and 2016, which included 36 studies [27], concluded that immigrants present a lower use rate of health services than the host populations. Among the reasons for not using these services are the lack of economic resources, language barrier, length of stay in the host country, and perceived discrimination [27].

The results of the present study suggest that Venezuelan immigrants with disabilities were more likely to not seek medical care when they became ill after arriving in Peru than Venezuelan immigrants without disabilities. Several studies have shown that people with disabilities have less access to health services [28,29]. A systematic review, which included 26 articles published between 2006 and 2016, concluded that people with disabilities have less access to preventive health services than the general population. This study identified that the main barriers to access were physical factors (non-accessible infrastructure), transportation (unavailability of adequate means of transportation, causing delays in getting to the appointment), negative attitude of health providers (perceived discrimination by persons with disabilities), and financial means (lack of money to pay for medical care) [30]. In addition, a study in Peru reported that older adults with disabilities were 63% less likely to have health insurance than those without disabilities [31].

Moreover, a study in the United States reported that people with disabilities and without insurance face more barriers to accessing health care than people without disabilities [12]. From the presented data, we can infer that Venezuelan immigrants with disabilities in Peru constitute a “doubly vulnerable” population group. The collective effect on education, work, or social relations is unknown. The lower use rate of health services by immigrants with disabilities should lead to efforts aimed at identifying the factors associated with this variable. One of the few studies on immigrants with disabilities in Latin America identified the lack of a disability card certifying their vulnerability condition as one of the main barriers to accessing health services [15]. Thus, it is necessary to implement a health insurance mechanism recognized in transit and host countries to avoid further health complications and prevent the aggravation of the sequelae of the disability of this doubly vulnerable population.

Moreover, the study of disability is complex because the results may vary according to the measurement methods employed. The evaluation of this condition among those with only one type of disability in comparison with those having two or more disabilities is not comparable. The same occurs when considering the levels of severity or the individual analysis of each type of disability, and variations even exist when assessing the time at which the disability occurred [32]. In addition, the use of different instruments increases this variation. As long as there is no widely accepted and applied consensus in population-based studies, the prevalence of disability will continue to differ between countries [33], especially in Latin America [34].

## Limitations and strengths

This study has the following limitations: First, the measurement of disability by self-perception could be conditioned by social desirability bias, which could provide less than accurate estimates of this condition. Second, failure to seek medical care could be related to other variables not available in the analyzed database. Third, it is not possible to determine whether the disability was acquired due to migration or before this event. Fourth, disability should be measured according to its levels of severity; however, since we did not have this information, we categorized this variable into a single type of disability and two or more types of disability. Fifth, the category of “two or more types of disability” was not considered in the results because of a few observations (n = 12). Finally, a cross-sectional survey cannot assume causality between the main variables. Despite these limitations, we consider that the findings are relevant because they represent the Venezuelan immigrant populations residing in Peru.

## Conclusions

In Peru, Venezuelan immigrants with a disability are more likely not to seek medical care than Venezuelan immigrants without a disability. The low prevalence of disability may be due to the “healthy migrant effect.” The prevalence of not seeking medical care among Venezuelan immigrants was lower than that among the general population. Based on these preliminary findings, we recommend conducting epidemiological studies to corroborate these results and identify the factors that influence the non-use of health services to improve the health conditions of this vulnerable population. We also suggest periodically quantifying the prevalence of disability in this population because, as a consequence of acculturation processes and cumulative disadvantage, these prevalence values will very likely increase to match the disability levels of the host population.

## Data Availability

All data produced are available online at https://proyectos.inei.gob.pe/microdatos/

https://proyectos.inei.gob.pe/microdatos/

## Data Availability

The datasets analyzed during the current study are available in the INEI web, [http://iinei.inei.gob.pe/microdatos]

## Conflicts of Interest

The authors have no relevant financial or non-financial interests to disclose.

## Funding Statement

No funding was received for conducting this study.

## Ethics approval

The project was reviewed and approved by the Institutional Research Ethics Committee of the Universidad Científica del Sur (Code: 197-2021-PREB15). The ENPOVE data are publicly accessible, do not allow the identification of participants, and are freely available to anyone who wishes to consult them.

## Authors’ contribution statements

All authors contributed to the study conception and design. Material preparation, data collection and analysis were performed by Mercedes Miranda-Tueros, Sonny Sthefanie Velarde-Meza and J. Jhonnel Alarco. The first draft of the manuscript was written by J. Jhonnel Alarco and all authors commented on previous versions of the manuscript. All authors read and approved the final manuscript.

## Notes

### Competing Interest Statement

The authors have declared no competing interest.

### Author Declarations

The datasets analyzed during the current study are available in the INEI web, [https://proyectos.inei.gob.pe/microdatos/]

